# COVID-19 vaccination breakthrough infections in a real-world setting: Using community reporters to evaluate vaccine effectiveness

**DOI:** 10.1101/2022.01.11.22268736

**Authors:** Matthew W Reynolds, Yiqiong Xie, Kendall B Knuth, Christina D Mack, Emma Brinkley, Stephen Toovey, Nancy A Dreyer

## Abstract

**Background:** COVID-19 has highlighted the need for new methods of pharmacovigilance. Here we use community volunteers to obtain systematic information on vaccine effectiveness and the nature and severity of breakthrough infections.

**Methods:** Between December 15, 2020 to September 16, 2021, 10,412 unpaid community-based participants reported the following information to an on-line registry: COVID-19 test results, vaccination (Pfizer, Moderna, or Johnson & Johnson), COVID-19 symptoms and perceived severity using a 4-point scale. COVID-19 infections were described for those who were 1) fully vaccinated, 2) partially vaccinated (received first of two dose vaccines or were <14 days post-final dose), or 3) unvaccinated.

**Results:** Of 8,554 who were vaccinated, COVID-19 infections were reported by 74 (1.0%) of those who were fully vaccinated and 198 (2.3%) of those who were partially vaccinated. Among the 74 participants who reported a breakthrough infection after full vaccination, the median time to reported positive test result was 104.5 days (Interquartile range: 77-135 days), with no difference among vaccine manufactures. One quarter (25.7%) of breakthrough infections in the fully vaccinated cases were asymptomatic. More than 97% of fully vaccinated participants reported no moderate/severe symptoms compared to 89.3% of the unvaccinated cases; and only 1.4% of fully vaccinated participants reported experiencing at least 3 moderate to severe symptoms compared to 7.8% in the unvaccinated.

**Conclusion:** Person-generated health data, also referred to as patient-reported outcomes, is a useful resource for quantifying breakthrough infections and their severity, showing here that fully vaccinated participants report no or very mild COVID-19 symptoms.

**Trial registration:** Clinicaltrials.gov NCT04368065, EU PAS Register EUPAS36240

## Background

The three COVID-19 vaccines authorized by the United States Food and Drug Administration (FDA) as of this writing have been found to be safe and highly efficacious in preventing symptomatic, laboratory-confirmed COVID-19 infections in randomized controlled trials.^1-3^ As is not uncommon with even the most effective vaccines, the currently authorized COVID-19 vaccines confer infection-permissive or ‘non-sterilizing’ immunity, i.e. while they may not always prevent infection, they will prevent or protect against illness, and particularly severe illness.^4^ Infection in the vaccinated is referred to as a ‘breakthrough’ infection; such cases are defined by the US Centers for Disease Control and Prevention (CDC) as those which have detectable SARS-CoV-2 RNA or antigen on a respiratory specimen collected ≥ 14 days after completion of a vaccine regimen.^1^ While cases of COVID-19 might still be expected to occur after vaccination, an excess of breakthrough infections could suggest a possible lack of vaccine effectiveness, consistent with the FDA definition of an adverse event as including “failure to produce the expected pharmacologic action”.^5^ One consideration of vaccine effectiveness would be the ability to reduce or prevent severe symptoms in those infected with SARS-CoV-2. Hence, while it is important to identify breakthrough cases, it is equally important to characterize the severity of the associated symptoms.

Understanding the safety of these new vaccines in the general population is of particular concern, both in terms of stemming the SARS-CoV-2 pandemic and of assuaging the vaccine hesitancy occasioned by the accelerated development of the currently available COVID-19 vaccines.^6,7^ Lack of drug effectiveness has been proven to be the most commonly reported adverse event (AE) in the US FDA Adverse Event Reporting System (FAERS),^6^ driven primarily by spontaneous adverse event reporting from consumers. From the pharmacovigilance perspective, assessing lack of drug efficacy can be challenging, with difficulties such as the definition of a simple or clear metric for an endpoint, and in this case, a lack of national systematic surveillance to detect infection. Such assessment is however especially relevant and important for these new COVID-19 vaccines which underwent accelerated evaluation and subsequently were rapidly deployed. Furthermore, the emergence of viral mutations conferring possible vaccine escape, including some known ‘variants of concern’, makes prompt detection of “lack of effectiveness” signals a public health imperative.^8^

While some serious safety concerns remain for these vaccines, e.g. anaphylaxis ^9,10^and Guillain-Barre Syndrome, ^11^ lack of vaccine effectiveness is of particular importance given the potential risk of severe COVID-19 disease and mortality. Lack of ability to effectively prevent, or to minimize, symptoms of infection, would imperil the success of current pandemic control efforts. Person-generated real-world data, sometimes called patient-reported health data or patient reported outcomes, should help to better understand and characterize breakthrough infections.

The objective of this work was to describe the pattern and characteristics of infections after vaccinations in recipients who received any of the three COVID-19 vaccines currently authorized for use in the U.S. (BioNTech (Pfizer), Moderna, Inc. (Moderna), and Janssen Pharmaceuticals (Johnson & Johnson (J&J)) or were never vaccinated. We describe the number of infections reported and patient-reported COVID-19 symptom information, stratified by vaccination status (fully vaccinated, partially vaccinated, and unvaccinated).

Additionally, we illustrate how data generated by health care recipients (in this case patients with confirmed COVID-19 infections) can be used for pharmacovigilance.

## Materials and methods

We leveraged data from a web-based registry, the COVID-19 Active Registry Experience (CARE, see www.helpstopcovid19.com). CARE was first launched on April 2, 2020 to study COVID-19 symptoms and severity outside of the hospital setting with the goal of understanding what factors, if any, could mitigate the impact of infection with COVID-19.^12-14^ The protocol and web technology are updated periodically, with the COVID-19 vaccination questions reported here added on December 15, 2020. Participants are recruited via social media, targeting adult U.S. residents. This study was approved by an Institutional Review Board and registered at both Clinicaltrials.gov NCT04368065 and the EU PAS register EUPAS36240. All participants provided informed consent online. The enrollment and follow-up surveys collect information on participants’ demographics, medical history, COVID-19-like symptoms and severity,^12^ COVID-19 tests, COVID-19 vaccination, and use of prescription and non-prescription medications and dietary supplements. Those who report having been vaccinated are also asked to provide the date of vaccination and side effects for first and second doses, as appropriate, manufacturer, and lot number, if known; participants are instructed to refer to their vaccination card to enhance data accuracy. Participants are asked to complete surveys once a week for the first month, and then twice a month for months two through twelve.

Surveys were submitted by 11,927 participants between December 15, 2020 and September 16, 2021. We excluded 101 reports of unauthorized vaccines, receipt of doses from more than one manufacturer, inconsistent dates, or vaccine dates earlier than June 2020. We also excluded participants who were unvaccinated and did not report a positive COVID-19 test (n=1,414). By doing this we attained participants who either received any FDA-authorized vaccination or reported a positive COVID-19. The analytic population consisted of 10,412 participants, including 8,554 who had ever received a Pfizer, Moderna, or J&J vaccine, and 1,858 who were never vaccinated but reported a positive COVID-19 test. Among the 8,554 participants, 7,532 completed vaccination (i.e., two doses of Pfizer or Moderna, or one dose of J&J), and 1,022 had one dose of Pfizer or Moderna. Vaccination status was determined at the time of their positive COVID-19 test (Figures 1a and 1b). For those who reported multiple positive COVID-19 test results, we considered tests within 90 days as the same infection and used the first positive test to mark the start of the infection. If there were positive tests more than 90 days apart, we used the first positive test reported within the latest (most current) course of infection. For participants who never received any vaccines during follow-up, we limited their positive tests to after December 15, 2020. Hence participants only had one test result selected for this analysis and one corresponding vaccination status at the time of that test result. There was no minimum requirement of follow-up time.

**Figure 1a.**
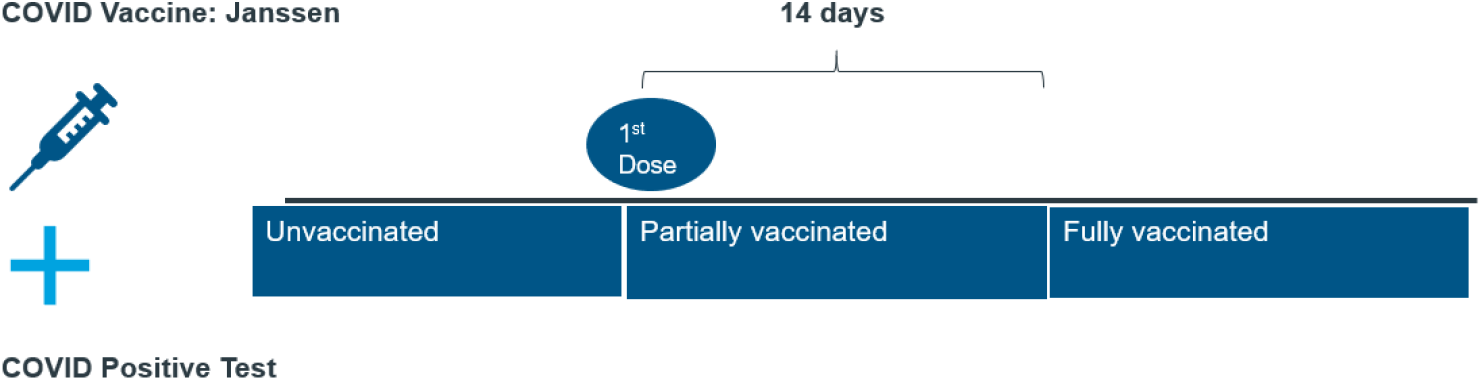
COVID-19 breakthrough infection journey: Janssen

**Figure 1b.**
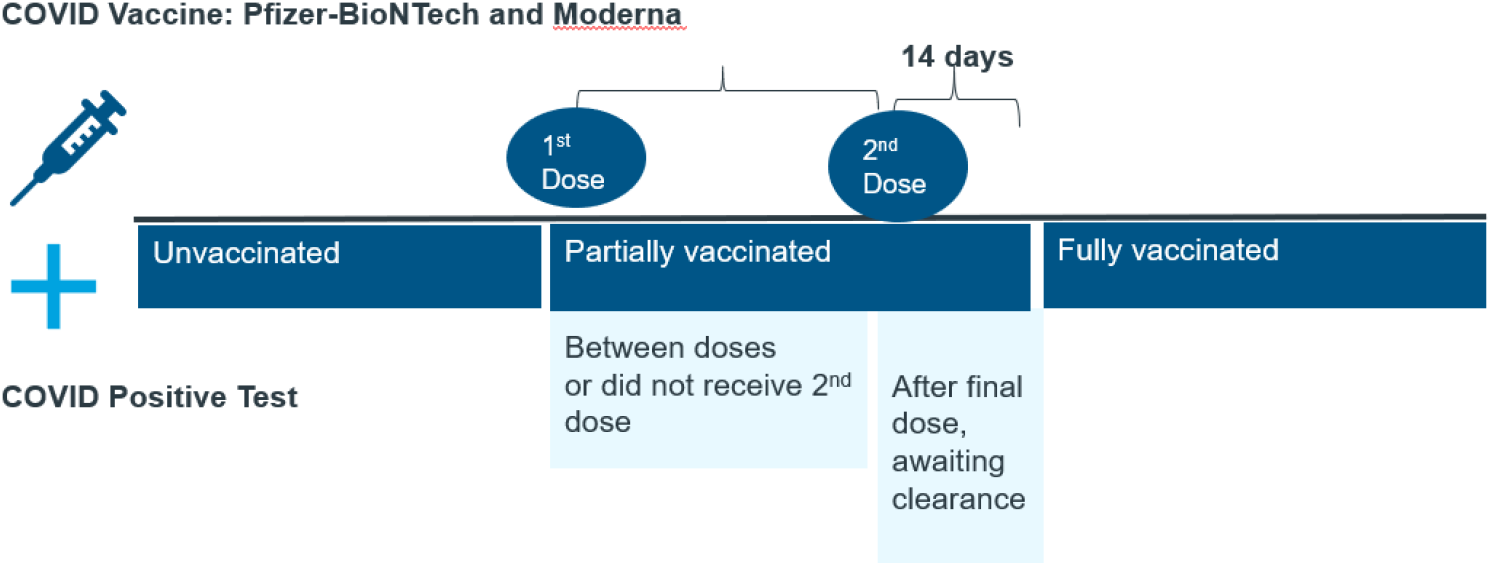
COVID-19 breakthrough infection journey: Pfizer-BioNTech and Moderna

## Results

### Characteristics of vaccinated study participants

There were 10,412 participants in our sample, including 8,554 who reported receiving at least one dose of an FDA-authorized vaccine and 1,858 who were never vaccinated but reported a positive COVID-19 test. The average age among the 8,554 vaccinated study participants was 47.9 years (range: 18-91 years); the majority were female (83.4%), with some college or higher education (90.8%), White (89.9%), non-smokers (90.4%), and received vaccination for influenza (73.7%) (Table 1). Among the 8,554 fully or partially vaccinated participants, 1,160 (13.6%) reported having an infection before any COVID-19 vaccination.

**Table 1.** Characteristics at enrollment of vaccinated participants

|  | All Participants<br>who received at<br>least 1 COVID-19 vaccine<br>N=8,554 | Vaccinated participants who later reported a<br>COVID-19 infection |  |
| --- | --- | --- | --- |
|  |  | Partially vaccinated at<br>time of infection<br>N=198 | Fully vaccinated at<br>time of infection<br>N=74 |
| Age in years (N) | 8553 | 198 | 74 |
| Mean (SD) | 47.9 (15.42) | 44.3 (13.03) | 44.5 (14.98) |
| Age Group (years, n[%]) | 8553 | 198 | 74 |
| 18-29 | 994 (11.6) | 25 (12.6) | 12 (16.2) |
| 30-39 | 2094 (24.5) | 59 (29.8) | 22 (29.7) |
| 40-49 | 1439 (16.8) | 43 (21.7) | 14 (18.9) |
| 50-59 | 1590 (18.6) | 40 (20.2) | 8 (10.8) |
| >=60 | 2436 (28.5) | 31 (15.7) | 18 (24.3) |
| Gender, n(%) | 8554 | 198 | 74 |
| Female | 7137 (83.4) | 170 (85.9) | 59 (79.7) |
| Male | 1200 (14.0) | 26 (13.1) | 13 (17.6) |
| Transgender/Other/ Not<br>Disclosed | 217(2.5) | 2(1) | 2(2.8) |
| Pregnant, n(%) | 7137 | 170 | 59 |
| Yes | 1019 (14.3) | 3 (1.8) | 12 (20.3) |
| Education, n(%) | 8538 | 198 | 74 |
| High school or less | 782 (9.2) | 21 (10.6) | 4 (5.4) |
| Some college | 2629 (30.8) | 70 (35.4) | 21 (28.4) |
| 4-yr college degree | 2212 (25.9) | 44 (22.2) | 19 (25.7) |
| >4yr college degree | 2915 (34.1) | 63 (31.8) | 30 (40.5) |
| Race, n(%) | 8514 | 198 | 74 |
| Black or African American | 149 (1.7) | 6 (3.0) | 3 (4.1) |
| White | 7680 (89.9) | 179 (90.4) | 68 (91.9) |
| Other | 712 (8.3) | 13 (6.6) | 3 (4.1) |
| Ethnicity, n(%) | 8491 | 195 | 74 |
| Hispanic or Latino | 553 (6.5) | 20 (10.3) | 5 (6.8) |
| BMI, n(%) | 8399 | 198 | 73 |
| Underweight or Normal<br>weight ($15.0 \leq \text{BMI} < 25.0$) | 2240 (26.7) | 36 (18.2) | 13 (17.8) |
|  |  | Partially vaccinated at<br>time of infection<br>N=198 | Fully vaccinated at<br>time of infection<br>N=74 |
| Overweight ($25.0 \leq \text{BMI} < 30.0$) | 2328 (27.7) | 53 (26.8) | 22 (30.1) |
| Obese ($30.0 \leq \text{BMI} \leq 40.0$) | 2820 (33.4) | 79 (39.9) | 28 (38.4) |
| Severe Obesity ($\text{BMI} > 40.0$) | 1011 (12.0) | 30 (15.2) | 10 (13.7) |
| Smoker, n(%) | 8217 | 197 | 73 |
| Yes | 789 (9.6) | 18 (9.1) | 8 (11.0) |
| Vaccinated for Influenza, n(%) | 8157 | 194 | 74 |
| Yes | 6012 (73.7) | 162 (83.5) | 58 (78.4) |
| Medical Conditions, n(%) | 8554 | 198 | 74 |
| Anxiety | 1459 (17.1) | 41 (20.7) | 12 (16.2) |
| Autoimmune disease | 593 (6.9) | 14 (7.1) | 6 (8.1) |
| Blood disorder | 69 (0.8) | 1 (0.5) | 0 |
| Cardiovascular disease | 300 (3.5) | 2 (1.0) | 3 (4.1) |
| Depression | 1742 (20.4) | 57 (28.8) | 12 (16.2) |
| Diabetes | 712 (8.3) | 13 (6.6) | 5 (6.8) |
| Hypertension | 1794 (21.0) | 51 (25.8) | 13 (17.6) |
| Insomnia or trouble sleeping | 512 (6.0) | 14 (7.1) | 3 (4.1) |
| Kidney disease | 31 (0.4) | 0 | 0 |
| Lung disease | 625 (7.3) | 14 (7.1) | 4 (5.4) |
| Organ transplant | 13 (0.2) | 0 | 0 |
| Whether on Treatment for<br>Cancer, n(%) | 8529 | 197 | 74 |
| On treatment | 99 (1.2) | 3 (1.5) | 1 (1.4) |

### COVID-19 infected study participants

The counts of participants who reported a positive COVID-19 test before receiving their first vaccination (n=1,160) and those who were never vaccinated and reported a positive COVID-19 test (n=1,858) are shown in Table 2. Among the 7,532 fully vaccinated participants, 74 (1.0%) participants reported an infection, a proportion that was generally similar across manufacturers with 6 participants (0.9%) for J&J, 52 participants (1.4%) for Pfizer, and 16 participants (0.5%) for Moderna. The proportion of participants who reported an infection during the period when they were partially vaccinated was 2.3% (n=198), also comparable across manufacturers with 0.2% for J&J, 2.4% for Pfizer, and 2.6% for Moderna.

**Table 2.** Number of participants with COVID-19 infections by vaccination status

|  | **Total** | **J & J** | **Pfizer** | **Moderna** | **Never vaccinated** |
| --- | --- | --- | --- | --- | --- |
| Total number of study participants | **11,826** | **652** | **4322** | **3580** | **3272** |
| Ever vaccinated | 8,554 | 652 | 4322 | 3580 |  |
| Completed vaccine regimen | 7,532 | 652 | 3788 | 3092 |  |
| Received only one dose of Pfizer/Moderna | 1022 |  | 534 | 488 |  |
| Never vaccinated | 3,272 |  |  |  | 3272 |
| **Total number of infected participants** | **3290** | **108** | **743** | **581** | **1858** |
| 1. Infection while fully vaccinated (i.e., after 14 days of last dose) *, n(%) | 74 (1.0) | 6 (0.9) | 52 (1.4) | 16 (0.5) |  |
| 2. Infected while partially vaccinated €, n(%) | 198 (2.3) | 1 (0.2) | 103 (2.4) | 94 (2.6) |  |
| After final dose, awaiting presumed immunity | 15 (0.2) | 1 (0.2) | 12 (0.3) | 2 (0.1) |  |
| Between two doses | 117 (1.7) |  | 64 (1.7) | 53 (1.7) |  |
| After 1st dose and did not receive 2nd dose | 66 (6.5) |  | 27 (5.1) | 39 (8.0) |  |
| 3. Infected while unvaccinated º, n(%) | 3018 (25.5) | 101 (15.5) | 588 (13.6) | 471 (13.2) | 1858 (56.8) |
| Infection prior to first dose | 1160 (13.6) | 101 (15.5) | 588 (13.6) | 471 (13.2) |  |
| Infection and never vaccinated | 1858 (56.8) |  |  |  | 1858 (56.8) |
| **Time interval from COVID-19 vaccination to infection** |  |  |  |  |  |
| **Infection after fully vaccinated (Days since presumed immunity) (N)** | 74 | 6 | 52 | 16 | N/A |
| Mean (SD) | 101.4 (44.99) | 78.7 (61.17) | 100.1 (42.82) | 113.9 (44.64) | N/A |
| Median [IQR] | 104.5 [77.0-135.0] | 91.5 [10.0-127.0] | 104.0 [75.0-132.5] | 108.5 [88.5-148.0] | N/A |
| Min - Max | 2-196 | 2-150 | 12-196 | 14-190 | N/A |
| **Partially vaccinated (Days since first dose) (N)** | 198 | 1 | 103 | 94 | N/A |
| Mean (SD) | 11.3 (21.84) | 8.0 (-) | 10.4 (14.24) | 12.4 (28.03) | N/A |
| Median [IQR] | 8.0 [5.0-12.0] | 8.0 [8.0-8.0] | 8.0 [5.0-13.0] | 8.0 [5.0-12.0] | N/A |
| Min - Max | 0-223 | 8-8 | 0-128 | 0-223 | N/A |

\* Percentages based on 7,532 participants who had completed doses
€ Percentages based on 8,554 participants who received any vaccination
º Percentages based on all 11,826 participants

Among the 74 participants who reported a breakthrough infection after full vaccination, the median time from 14 days post-final vaccine to reported positive test result was 104.5 days, Interquartile range (IQR): 77-135 days. There were no significant differences across vaccine manufactures, with 91.5 days for J&J, 104.0 days for Pfizer, and 108.5 days for Moderna (p-value = 0.44). Among the 74 people infected after full vaccination, their average age was 44.5 years. Most were female (79.7%), White (91.9%), and 40.5% had above a college degree (Table 3). Among the 198 people infected after partial vaccination, median time from the first dose to infection was 8.0 days (IQR: 5.0-12.0 days).

**Table 3.** Characteristics at survey enrollment of COVID-19 positive participants

|  | All COVID-19<br>Infections<br>N=3290 (%) | Unvaccinated*<br>N=3018 (%) | Partially<br>Vaccinated<br>N=198 (%) | Fully<br>vaccinated<br>N=74 (%) |
| --- | --- | --- | --- | --- |
| Age in years (N) | 3290 | 3018 | 198 | 74 |
| Mean (SD) | 42.3 (13.40) | 42.2 (13.37) | 44.3 (13.03) | 44.5 (14.98) |
| Age Group (years, n[%]) | 3290 | 3018 | 198 | 74 |
| 18-29 | 594 (18.1) | 557 (18.5) | 25 (12.6) | 12 (16.2) |
| 30-39 | 907 (27.6) | 826 (27.4) | 59 (29.8) | 22 (29.7) |
| 40-49 | 784 (23.8) | 727 (24.1) | 43 (21.7) | 14 (18.9) |
| 50-59 | 587 (17.8) | 539 (17.9) | 40 (20.2) | 8 (10.8) |
| >=60 | 418 (12.7) | 369 (12.2) | 31 (15.7) | 18 (24.3) |
| Gender, n(%) | 3290 | 3018 | 198 | 74 |
| Female | 2821 (85.7) | 2592 (85.9) | 170 (85.9) | 59 (79.7) |
| Male | 434 (13.2) | 395 (13.1) | 26 (13.1) | 13 (17.6) |
| Transgender/Other/ Not<br>Disclosed | 35(1.1) | 31(1.0) | 2(1) | 2(2.7) |
| Pregnant, n(%) | 2821 | 2592 | 170 | 59 |
| Yes | 73 (2.6) | 58 (2.2) | 3 (1.8) | 12 (20.3) |
| Education, n(%) | 3289 | 3017 | 198 | 74 |
| High school or less | 564 (17.2) | 539 (17.9) | 21 (10.6) | 4 (5.4) |
| Some college | 1304 (39.7) | 1213 (40.2) | 70 (35.4) | 21 (28.4) |
| 4-yr college degree | 715 (21.7) | 652 (21.6) | 44 (22.2) | 19 (25.7) |
| >4yr college degree | 706 (21.5) | 613 (20.3) | 63 (31.8) | 30 (40.5) |
| Race, n(%) | 3286 | 3014 | 198 | 74 |
| Black or African American | 202 (6.2) | 193 (6.4) | 6 (3.0) | 3 (4.1) |
| White | 2640 (80.3) | 2393 (79.4) | 179 (90.4) | 68 (91.9) |
| Other | 444 (13.5) | 428 (14.2) | 13 (6.6) | 3 (4.1) |
| Ethnicity | 3245 | 2976 | 195 | 74 |
| Hispanic or Latino | 538 (16.6) | 513 (17.2) | 20 (10.3) | 5 (6.8) |
| BMI, n(%) | 3242 | 2971 | 198 | 73 |
| Underweight or Normal<br>weight ($15.0 \leq \text{BMI} < 25.0$) | 795 (24.5) | 746 (25.1) | 36 (18.2) | 13 (17.8) |
| Overweight ($25.0 \leq \text{BMI} < 30.0$) | 825 (25.5) | 750 (25.2) | 53 (26.8) | 22 (30.1) |
| Obese ($30.0 \leq \text{BMI} \leq 40.0$) | 1145 (35.2) | 1038 (34.9) | 79 (39.9) | 28 (38.4) |
| Severe Obesity (BMI>40.0) | 477 (14.7) | 437 (14.7) | 30 (15.2) | 10 (13.7) |
| Smoking status, n(%) | 3211 | 2941 | 197 | 73 |
| Smoker | 342 (10.7) | 316 (10.7) | 18 (9.1) | 8 (11.0) |
| Vaccination status for<br>Influenza, n(%) | 3231 | 2963 | 194 | 74 |
| Vaccinated | 1867 (57.8) | 1647 (55.6) | 162 (83.5) | 58 (78.4) |
| Medical Conditions, n(%) | 3290 | 3018 | 198 | 74 |
| Anxiety | 608 (18.5) | 555 (18.4) | 41 (20.7) | 12 (16.2) |
| Autoimmune disease | 192 (5.8) | 172 (5.7) | 14 (7.1) | 6 (8.1) |
| Blood disorder | 10 (0.3) | 9 (0.3) | 1 (0.5) | 0 |
| Cardiovascular disease | 19 (0.6) | 14 (0.5) | 2 (1.0) | 3 (4.1) |
| Depression | 594 (18.1) | 525 (17.4) | 57 (28.8) | 12 (16.2) |
| Diabetes | 238 (7.2) | 220 (7.3) | 13 (6.6) | 5 (6.8) |
| Hypertension | 634 (19.3) | 570 (18.9) | 51 (25.8) | 13 (17.6) |
| Insomnia or trouble sleeping | 210 (6.4) | 193 (6.4) | 14 (7.1) | 3 (4.1) |
| Kidney disease | 4 (0.1) | 4 (0.1) | 0 | 0 |
| Lung disease | 221 (6.7) | 203 (6.7) | 14 (7.1) | 4 (5.4) |
| Organ transplant | 1 (0.03) | 1 (0.03) | 0 | 0 |
| Whether on Treatment for<br>Cancer, n(%) | 3272 | 3001 | 197 | 74 |
| On treatment | 29 (0.9) | 25 (0.8) | 3 (1.5) | 1 (1.4) |

\*Unvaccinated group included participants reporting infection before any vaccination and those reporting infection and never vaccinated

There were small differences in age across the three vaccination status groups, with 24.3% of people aged ≥ 60 years in the fully vaccinated group and 15.7% and 12.2% in partially vaccinated and unvaccinated, respectively. Partially vaccinated and unvaccinated groups also had lower proportion of males (13.1% and 13.1% compared with 17.6% among fully vaccinated) and above college education (31.8% and 20.3%, compared with 40.5% among fully vaccinated). Other demographics and medical characteristics appeared to be similar across the three subgroups.

### COVID-19 symptoms and impact among vaccinated vs. unvaccinated

When examining COVID-19 symptoms associated with a reported positive COVID-19 test, the fully vaccinated group had lower proportions of every COVID-19 symptom compared to the partially vaccinated and unvaccinated groups, including headache, aches and pains, decreased sense of taste and smell, chills, and diarrhea (Table 4 and Figure 2). Fully vaccinated breakthrough cases reported fewer mean number of COVID-19 symptoms (4.1) when compared to partially vaccinated (5.8) or unvaccinated cases (5.3).

**Table 4.** COVID-19 infection symptoms among COVID-19 positive participants

|  | All COVID-19<br>Infections<br>N=3,290 (%) | Unvaccinate<br>d*<br>N=3,018 (%) | Partially<br>Vaccinated<br>N=198 (%) | Fully vaccinated<br>N=74 (%) |
| --- | --- | --- | --- | --- |
| **COVID-19 Symptoms, n(%)** |  |  |  |  |
| Aches and Pains | 1,381 (42.0) | 1287 (42.6) | 76 (38.4) | 18 (24.3) |
| Chills | 850 (25.8) | 780 (25.8) | 58 (29.3) | 12 (16.2) |
| Decreased Sense of Smell | 1469 (44.7) | 1362 (45.1) | 84 (42.4) | 23 (31.1) |
| Cough | 1606 (48.8) | 1467 (48.6) | 108 (54.6) | 31 (41.9) |
| Decreased Sense of Taste | 1349 (41.0) | 1247 (41.3) | 77 (38.9) | 25 (33.8) |
| Diarrhea | 784 (23.8) | 726 (24.1) | 48 (24.2) | 10 (13.5) |
| Fatigue | 2,051 (62.3) | 1871 (62.0) | 136 (68.7) | 44 (59.5) |
| Fever | 680 (20.7) | 626 (20.7) | 46 (23.2) | 8 (10.8) |
| Nausea | 712 (21.6) | 656 (21.7) | 44 (22.2) | 12 (16.2) |
| Nasal Congestion | 1,677 (51.0) | 1,540 (51.0) | 105 (53.0) | 32 (43.2) |
| Runny Nose | 1,054 (32.0) | 961 (31.8) | 73 (36.9) | 20 (27.0) |
| Shortness of Breath | 906 (27.5) | 837 (27.7) | 53 (26.8) | 16 (21.6) |
| Sore Throat | 906 (27.5) | 820 (27.2) | 67 (33.8) | 19 (25.7) |
| Vomiting | 164 (5.0) | 150 (5.0) | 12 (6.1) | 2 (2.7) |
| Headache | 1744 (53.0) | 1596 (52.9) | 117 (59.1) | 31 (41.9) |
| **Number of symptoms** |  |  |  |  |
| Mean (SD) | 5.3 (3.87) | 5.3 (3.87) | 5.8 (3.90) | 4.1 (3.73) |
| Median [IQR] | 5.0 [2.0-8.0] | 5.0 [2.0-8.0] | 5.0 [2.0-9.0] | 4.0 [0.0-6.0] |
| Min-Max | 0-15 | 0-15 | 0-15 | 0-14 |
| **Patients with number of symptoms, n(%)** |  |  |  |  |
| 0 symptoms | 542 (16.5) | 496 (16.4) | 27 (13.6) | 19 (25.7) |
| 1-2 symptoms | 418 (12.7) | 381 (12.6) | 27 (13.6) | 10 (13.5) |
| 3+ symptoms | 2330 (70.8) | 2141 (70.9) | 144 (72.7) | 45 (60.8) |
| **Patients with number of moderate or severe symptoms, n(%)** |  |  |  |  |
| 0 moderate or severe symptoms | 2945 (89.7) | 2691 (89.3) | 182 (92.4) | 72 (97.3) |
| 1-2 moderate or severe symptoms | 91 (2.8) | 87 (2.9) | 3 (1.5) | 1 (1.4) |
| 3+ moderate or severe symptoms | 247 (7.5) | 234 (7.8) | 12 (6.1) | 1 (1.4) |
| **Hospitalization** |  |  |  |  |
| Hospitalized | 54(1.6) | 49(1.6) | 4(2) | 1(1.4) |

\*Unvaccinated group included participants reporting infection before any vaccination and those reporting infection and never vaccinated

**Figure 2.**
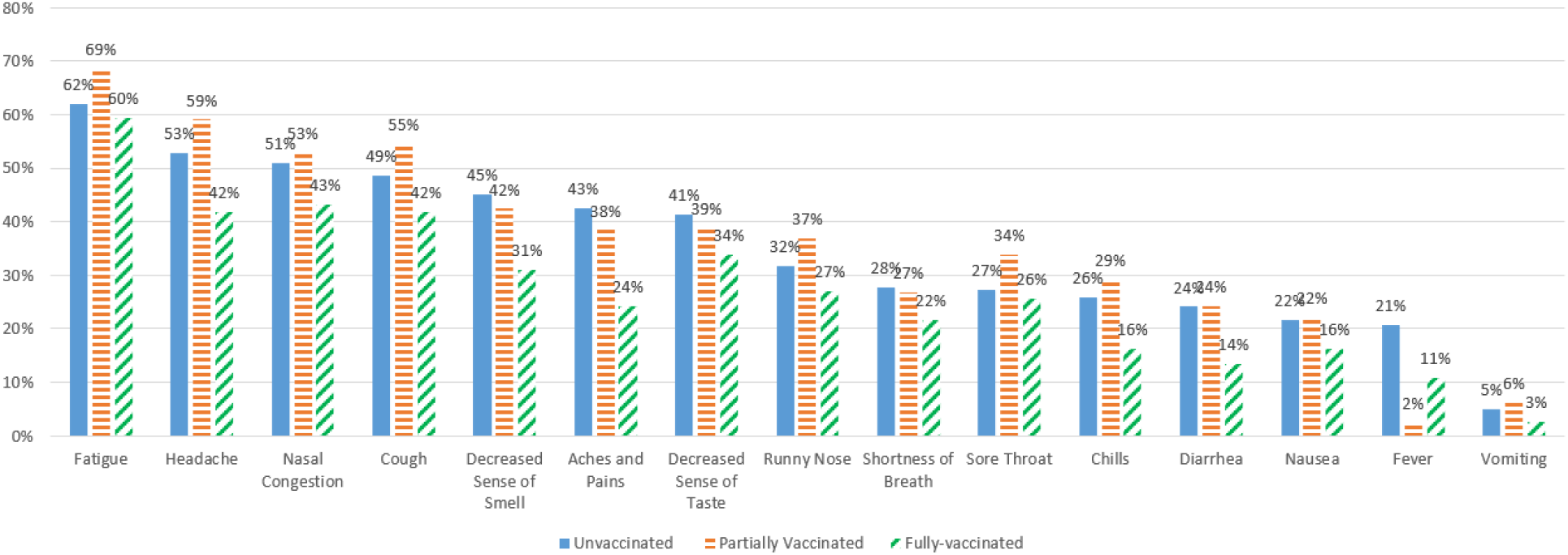
Symptoms reported by vaccination status

Approximately one quarter (25.7%) of fully vaccinated breakthrough cases reported no symptoms compared with 13.6% and 16.4% among partially vaccinated and unvaccinated cases, respectively (Figure 3). Fully vaccinated breakthrough cases were also more likely to report only mild symptoms and not report moderate or severe symptoms (97.3%) compared with partially vaccinated or unvaccinated cases (92.4% and 89.3%, respectively) (Table 4 and Figure 4).

**Figure 3.**
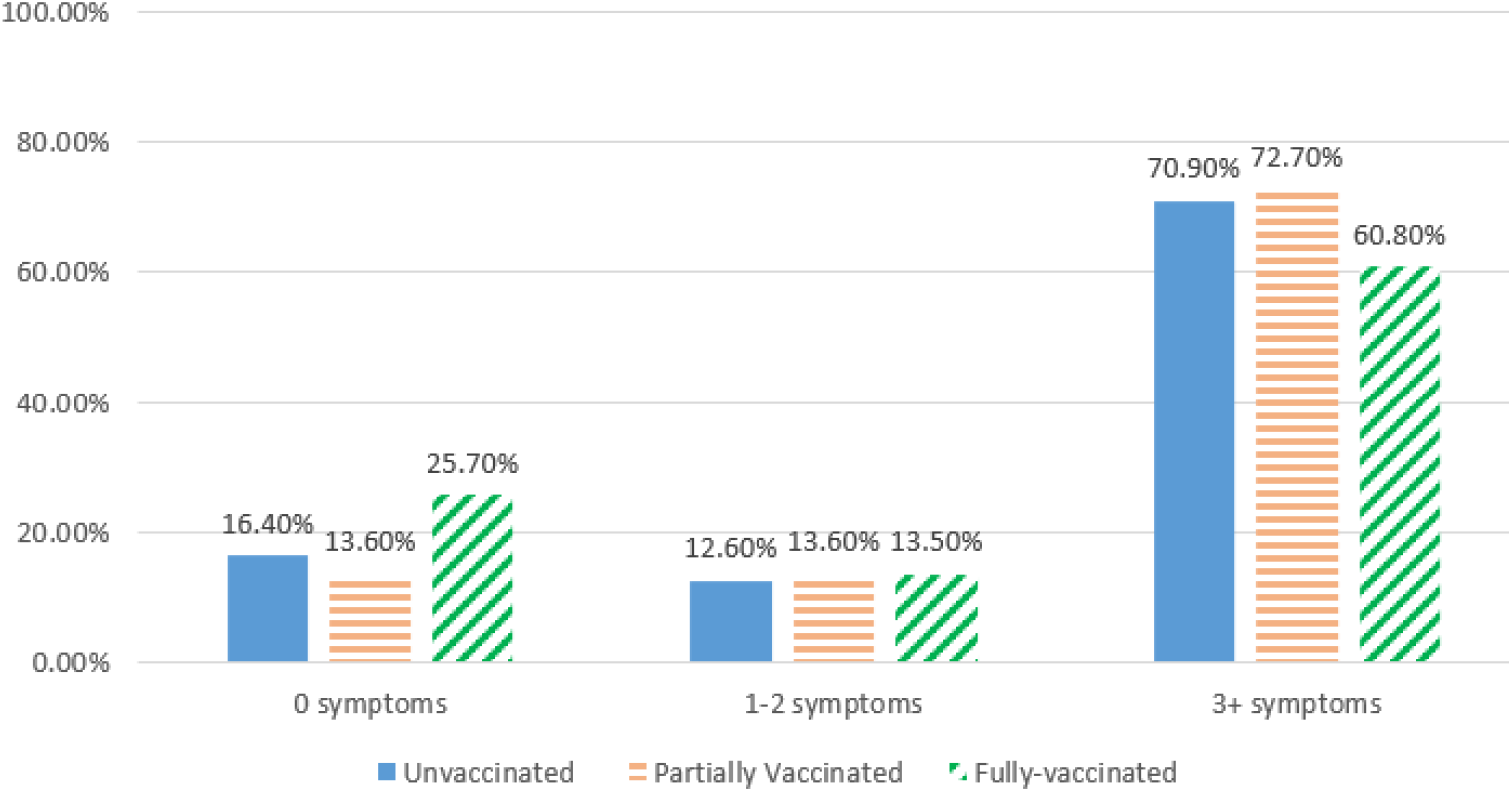
Number of symptoms reported by vaccination status

**Figure 4.**
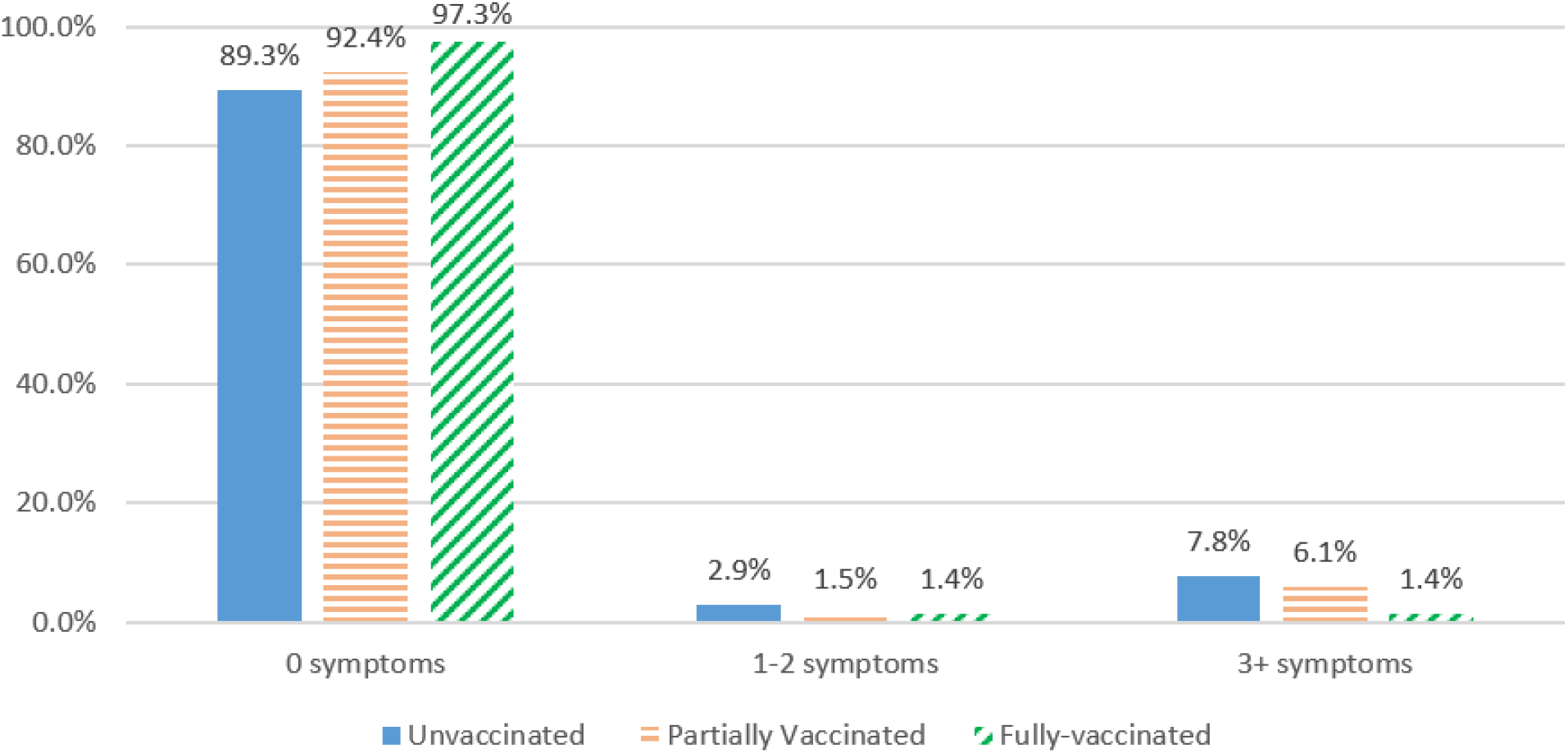
Number of moderate or severe symptoms reported by vaccination status

## Discussion

Systematic collection of person-generated health data as illustrated by the CARE registry demonstrates the value of coupling symptom information with COVID-19 test data to provide meaningful and timely information about vaccine effectiveness. This study provides quantitative estimates of whether and how intensely cases of COVID-19 occur in people who are fully vaccinated, a question of importance given the continuing emergence of variants of concern.^15^ By far the majority of the infections reported in this study occurred before day 14 after recipient’s final dose, noting that a relatively limited follow-up time precludes investigation of wanning immunity. Our findings confirm earlier reports.^1,16^

This study provides confirmatory evidence that vaccination, although ‘non-sterilizing’ or ‘infection-permissive’, affords significant protection against severe symptoms of COVID-19 disease, and suggests that it also confers protection against infection overall. In this cohort, 0.9% of all participants reported infections after full vaccination, which is consistent with the low rates reported elsewhere.^17^ Strikingly, approximately 25% of infections reported by fully vaccinated individuals were asymptomatic. Moreover, fully vaccinated individuals who experienced breakthrough infections reported significantly fewer symptoms than partially vaccinated or unvaccinated cases, with only a small fraction of fully vaccinated cases reporting moderate or severe symptoms and about one quarter experienced no symptoms at all. Thus, our findings are consistent with many studies reporting that only a small proportion of breakthrough cases are severe, and that a significant proportion of breakthrough infections are mild or asymptomatic.^16,18-21^

In this study, there were a few differences between the participants who reported COVID-19 infection after full vaccination and those who reported infection while partially vaccinated. The fully vaccinated group had a slightly higher proportion of infection in males, individuals aged 60 years or above and those with at least a college education. Reasons for the differences are not clear; potential explanations include that these participants (older and more educated) may have been vaccinated earlier, and thus have had a longer time after being fully vaccinated; such individuals would have had a longer ‘at risk’ time and/or potentially waning immunity. Older age (>60 years) might also reflect a degree of immunosenescence and a less robust response to vaccination.^22^ It should be noted that several factors that might be expected to be associated with an increased risk of infection such as hypertension, diabetes, and obesity, were not associated with COVID-19 infection in the fully vaccinated. This observation might offer encouraging mechanistic support for vaccine effectiveness but does leave open potential explanations for the occurrence of breakthrough.

Also of interest, among the fully vaccinated participants, the median time to breakthrough infection is 105 days, ranging from 2 to 196 days. Other recent studies have found similar time to breakthrough cases, including a study in Massachusetts that found a median time of 86 days after final vaccine dose. ^18,23^ Interestingly, in this study, the average time to breakthrough infection by manufacturer was not significantly different with 91.5, 104.0, and 108.5 days for J&J, Pfizer, and Moderna, respectively. Among the 198 people infected after partial vaccination, the median time to infection was 8.0 days; while this average time is much shorter than those who were fully vaccinated, it should be considered that the window for having infection while partially vaccinated was by definition confined to being shorter with a window of only 14 days until full vaccination for J&J and about 28 days between doses plus 14 days post-2^nd^ dose (42 days) for those receiving Pfizer or Moderna.

The proportions of participants that reported infections after being fully vaccinated were not high, but they were notably higher (1.4%) for participants vaccinated with Pfizer as compared to J&J (0.9%) and Moderna (0.5%); The proportion of participants reporting infection while partially vaccinated were similar for those vaccinated with Pfizer vaccine (2.4%) and Moderna (2.6%), but lower for those vaccinated for J&J (0.2%). This result is not surprising as noted previously, as the potential time window for full vaccination for those receiving J&J is by definition much shorter (approximately a third as long) than those receiving Pfizer or Moderna.

This study has a number of strengths, particularly the comparison of COVID symptoms between vaccinated and unvaccinated cases. This rich symptom data permits further contextualization of the effectiveness of the vaccine beyond simply reporting a positive or negative COVID-19 test result. Vaccine reporting is likely to be more complete than the vaccination status recorded in most real-world data sources such as insurance claims data and/or electronic health record datasets, considering how vaccines were distributed in the United States and the lack of any single accessible central source of vaccination status information for individual patients. Similarly, COVID-19 test results are also likely to be more complete than in some existing real-world databases, since participants were systematically queried to see if they were tested at every bi-weekly follow-up. Furthermore, these data were collected and able to be analyzed in near real-time, thus providing an opportunity to conduct pharmacovigilance activities rapidly and allowing for immediate opportunity to intervene if and when merited.

Nonetheless it is important to keep in mind that, like other online registries that survey volunteers without intervention or collaboration from medical care providers, the CARE registry relies on participants to contribute information completely, faithfully and accurately. Missing data, especially outcomes of interest, are possible and these data were not validated with respect to documenting the COVID-19 tests or obtaining a clinical assessment of the person-reported symptoms. Nevertheless, the validity of reports from lay people on medication use has been demonstrated,^1^ and as such, reporting quality is likely to apply at least as well to vaccine reporting. Using a direct to patient/person approach for future pharmacovigilance activities could support incorporation of exposure or outcome validation via direct to patient/physician follow-up and/or linkage to patient medical records.

There also may be underreporting of breakthrough cases and potential misclassification of their timing and severity. Participation in follow-up surveys depends on sustained personal interest, and participants may have tested positive after reporting their vaccine information but not completed any follow-up surveys. It is also possible that individuals with asymptomatic breakthrough infections were not aware of their infection in the absence of widespread systematic surveillance testing in most parts of the US. Very serious cases of COVID-19 that required hospitalization or resulted in death may be underreported. Additionally, participants who were vaccinated against COVID-19 may be less likely to be tested, especially if asymptomatic. Analytically, if only one positive test was reported, we assumed the positive test date reflected the start of the infection, whereas the infection may have started earlier and gone undetected prior to vaccination initiation.

Although CARE is not a representative, systematic sample of the U.S. adult population, it has broad geographic representation, with participants from all 50 states. Respondents include those with internet access and sufficient availability and interest to participate in surveys. Furthermore, projects like this tend to attract more females as well as those “worried well,” with higher participation among those with autoimmune disorders as well as anxiety, depression and insomnia. More importantly, it is unlikely that there was differential participation by vaccine manufacturers. Therefore comparisons between vaccine manufacturers in the CARE population are not likely to be biased.

## Conclusion

The systematic collection of person-generated health data shows that only a small proportion of vaccinated people become infected after vaccination, and they are more likely to be asymptomatic or have fewer and milder symptoms than unvaccinated people. This insight supports the evidence that COVID-19 vaccines are effective in the real world and that while cases of COVID-19 may occur in vaccinated individuals, these are uncommon, and generally mild in nature. The vaccines appear to be effective at preventing severe disease. This efficient direct-to-patient approach can obtain information not readily available in most real world data sources, and can be used effectively to study adverse events including product effectiveness.

## Data Availability

Due to data privacy and security regulations the researchers are not able to share participant level data.

## Acknowledgements

We thank Lisa Albert for her work on preliminary analyses, Savitha Pallipuram for leading the Technical team, and Alex Secora for his thoughtful editing contributions.

